# A Delphi process to determine clinicians’ attitudes and beliefs towards paediatric major incident triage within the United Kingdom

**DOI:** 10.1101/2022.02.09.22270720

**Authors:** J Vassallo, S Blakey, P Cowburn, J Surridge, JE Smith, B Scholefield, Mark D Lyttle, PERUKI (Paediatric Emergency Research in the UK and Ireland)

**Affiliations:** Institute of Naval Medicine, Gosport, PO12 2DL, UK; Academic Department of Military Emergency Medicine, Royal Centre for Defence Medicine, Birmingham, UK; Great Western Air Ambulance Charity, Almondsbury, Bristol, UK; Emergency Department, Bristol Royal Hospital for Children, Bristol UK; Research in Emergency Care, Avon Collaborative Hub (REACH), University of the West of England, Bristol, UK; Emergency Department, University Hospitals Bristol and Weston NHS Foundation Trust, Bristol, UK; South Western Ambulance Service NHS Foundation Trust, North Bristol Operations Centre, Bristol, UK; National Ambulance Resilience Unit (NARU), College of Policing, Ryton on Dunsmore, UK; Emergency Department, University Hospitals of Derby and Burton NHS Foundation Trust, Derby, UK; Emergency department, University Hospitals Plymouth NHS Trust, Plymouth, UK; Birmingham Acute Care Research Group, Institute of Inflammation and Ageing, University of Birmingham, UK; Paediatric Intensive Care, Birmingham Women & Children’s Hospital, NHS Foundation Trust, Birmingham, UK

**Author notes:** **Corresponding author:** Dr J Vassallo. Institute of Naval Medicine, Alverstoke, Gosport, PO12 2DL, UK. **Funding** No external funding was received for this study. **Author Contributions** JV and ML conceived the study idea, with survey content designed by JV, JES, JS, PC, BS, ML. The study was conducted by JV and ML. JV, SB, ML conducted the analysis and interpretation. SB wrote the initial draft of the manuscript. All authors contributed with critical revisions to the manuscript and approved the final manuscript. JV takes responsibility for the manuscript as a whole.

**Keywords:** Triage, paediatrics, major incidents, life-saving interventions

## Abstract

**Introduction:** Triage is a key principle in the effective management of major incidents, yet there is a paucity of evidence surrounding the optimal method of paediatric major incident triage (MIT). This study aimed to derive consensus on key components of paediatric MIT among healthcare professionals involved in the management of paediatric major incidents.

**Methods:** This modified two-round online Delphi consensus study, delivered between July and October 2021, included participants from pre-hospital and hospital specialities involved in managing a paediatric major incident. Statements were derived iteratively based on review of MIT tools, and extant literature. A 5-point Likert agreement scale was used to determine consensus, which was set a priori at 70%.

**Results:** 111 clinicians completed both rounds, with 13 of 17 statements reaching consensus. Positive consensus was reached on the use of rescue breaths in mechanisms associated with hypoxia or asphyxiation, use of mobility assessment as a crude discriminator of injury, and use of adult physiology for older children. Whilst positive consensus was reached on the benefits of a single MIT tool for use across the entire adult and paediatric age range, there was negative consensus in relation to the clinical implementation of such a tool. Consensus could not be reached regarding the use of a single tool across the whole paediatric age range specifically, nor on the use of rescue breaths in blunt or penetrating trauma.

**Conclusion:** This Delphi study has established consensus among a large group of subject matter experts on several key elements of paediatric MIT. Further work is required to develop a triage tool that can be implemented based on emerging and ongoing research, and which is acceptable to clinicians.

**What this paper adds?:** *Section 1: What is already known on this subject?:*

∘ Whilst triage is a key principle in the effective management of a major incident, there is limited evidence surrounding the use of existing paediatric major incident triage (MIT) tools
∘ Paediatric MIT tools currently used in the UK are associated with high rates of under-triage, failing to identify those in need of life-saving interventions
∘ Existing paediatric MIT tools differ from adult tools, including approach to physiological ranges, and recommendation for initiation of rescue breaths

*Section 2: What this study adds:*

∘ Consensus was reached supporting use of rescue breaths for mechanisms associated with hypoxia or asphyxiation, mobility as a crude discriminator for serious injury, and adult physiology for older children
∘ Whilst consensus was reached on benefits related to use of a single tool across all age ranges (adult and paediatric), the expert panel did not support this approach for actual clinical practice
∘ There was no consensus on use of rescue breaths in blunt or penetrating trauma, or use of a single triage tool for the entire paediatric age group
∘ Further work is required to develop and implement a MIT tool that accurately identifies children needing life-saving interventions, and that is acceptable to clinicians

## BACKGROUND

Major incidents occur worldwide on a regular basis and are characterised by the need for additional resources with which to respond to the incident; furthermore, they have the potential to overwhelm healthcare resources and cause patient harm.^1,2^ Major Incident Triage (MIT) tools aim to mitigate these risks by prioritising patients with high clinical acuity and need for life-saving intervention, whilst also identifying a cohort at lower risk.^1^ Major incidents involving children are less common,^3^ but they also carry potential for long-lasting negative psychological effects on clinicians.^4^ The ideal paediatric MIT tool should therefore be simple and rapid to apply, demonstrate good performance accuracy, and provide decision making support to clinicians.^5^

Whilst a number of MIT tools are used internationally,^6^ the Modified Physiological Triage Tool-24 (MPTT-24) has recently been implemented as the MIT tool of choice for adults in UK in-hospital practice due to its performance accuracy.^2,7^ The two most widely used paediatric MIT tools in the UK are JumpSTART ^8^ for hospital settings, and the Paediatric Triage Tape (PTT)^9^ for the pre-hospital setting. The MPTT-24 (and a paediatric version) have recently been found to outperform existing paediatric MIT tools on a trauma registry dataset, in their ability to identify patients in need of a life-saving intervention.^10,11^ However, limitations of existing evidence, patient and professional factors, and existing life support guidance^12^ cloud direct translation of these findings to practice.

Although the MPTT-24 incorporates physiological variables, translation into paediatric clinical practice is more complex as normal ranges change with age. The consequent potential for operator error has led to suggestions that physiological variables should be removed from paediatric MIT tools, or that adult variables should be used for adolescents. Whilst rescue breaths are recommended in resuscitation guidance for individual children^12^ and in JumpSTART, this is likely to delay the assessment of subsequent patients when faced with multiple casualties. Mortality and need for life-saving interventions are both highest in the youngest age cohort,^10^ yet these patients are the most difficult to rapidly assess. For example, mobility assessment is a key factor in several MIT tools, but often does not account for young children being developmentally pre-mobile. Some have therefore suggested that all patients below a lower age threshold should be assigned the highest priority (Priority 1; P1).

The aim of this study was to seek consensus opinion of healthcare professionals involved in the triage and management of paediatric patients at major incidents, determining the level of agreement with existing practice and concepts arising from emerging evidence in order to determine the optimum manner in which paediatric MIT should be conducted.

## METHODS

An online modified Delphi consensus study,^13,14^ consisting of two rounds, was undertaken between July and October 2021. Round 1 ran for 4 weeks (23/07/2021-20/08/2021) and Round 2 for 3 weeks (15/09/2021-06/10/2021), with a maximum of three reminders sent in each round. Surveys were delivered using Research Electronic Data Capture tools (REDCap),^15,16^ a secure, web-based software platform. Data were stored securely on a server within University Hospitals Bristol and Weston NHS Foundation Trust.

Potential participants were identified by accessing Subject Matter Expert groups across the UK and Ireland including the National Ambulance Services Medical Directors Group (NASMeD), Faculty of Pre-Hospital Care (FPHC), Paediatric Emergency Research in the UK & Ireland (PERUKI)^17^ and the Paediatric Critical Care Society Study Group (PCCS-SG). Participants could also invite other relevant subject matter experts to participate. Participants were invited by email or social media, using a link which included relevant background study information. This included existing and emerging research, example scenarios, and MIT tools used in children. The invitation highlighted the need for participation in both rounds, anticipated time for completion, and the link to the consensus statements. Those who did not complete Round 1 were ineligible to complete the subsequent round, and no new invitations were sent after completion of Round 1. Participants could exit either round, at any stage, if they no longer wished to continue. Participants were identifiable through their email address, but these were only available to the data manager, and all data extracted for analysis were anonymised.

Consensus statements were developed iteratively by the study team following a review of relevant literature, and adult and paediatric MIT tools. Statements were grouped in categories, with supplementary information provided for each topic area: (i) rescue breaths, (ii) mobility assessment, (iii) physiological variables and age ranges, (iv) older paediatric patients, and (v) very young children. A five-point Likert scale, from strongly disagree to strongly agree, was used for categorical statements. For statements seeking opinion on continuous variables (for example, upper or lower age thresholds), a range of options was presented using a drop-down list. Consensus was agreed a priori and set at 70% (either agree/strongly agree or disagree/strongly disagree).

Round 1 consisted of 16 statements; any that reached the pre-specified level for consensus were removed in Round 2. Statements which did not reach consensus in Round 1 were reviewed in line with respondent feedback, and either repeated verbatim or modified for Round 2 where necessary. Prior to Round 2, respondents were sent summary results and their individual responses from Round 1, and a document providing the rationale for any changes to statements in Round 2. One question was modified for Round 2, differentiating the use of rescue breaths into cohorts of blunt and penetrating trauma.

Data were analysed anonymously and following each round, responses were grouped into three categories consisting of “Agree” (agree/strongly agree), “Neither Agree/Disagree” and “Disagree” (disagree/strongly disagree). Incomplete responses were excluded in each round; however, if a participant’s response was incomplete only for Round 2, their responses to Round 1 were included. Consensus agreement is presented as proportions for both categorical and continuous variables; medians and interquartile ranges are presented using the 5-point Likert to show spread of opinion. Analyses were conducted in Microsoft Excel (Version 2016).

### Ethics

As this was a survey of health professionals identified via existing collaborative networks, formal ethics approval was not required according to the HRA framework decision tool.^18^

## RESULTS

In total 157 participants completed Round 1, of which 111 (70.7%) provided complete responses in Round 2. Participants were predominantly from a pre-hospital (47%) or Paediatric Emergency Medicine (PEM; 41%) background, with 63.7% being of consultant grade and 24.2% either paramedics or extended-scope paramedics. Specialities and grades of participants are presented in Table 1.

**Table 1:** Demographics of study participants.

| Speciality* | Grade; number of complete responses, separated by speciality |  |  |  |  |  |  |  |
| --- | --- | --- | --- | --- | --- | --- | --- | --- |
|  | Consultant | ST | AS | ACP | Paramedic | ES-Paramedic | Nurse | Total |
| **Round 1** |  |  |  |  |  |  |  |  |
| Pre-hospital | 27 | 7 | 1 | 1 | 21 | 16 | 1 | 74 |
| EM | 25 | 5 | 2 | 2 | 0 | 0 | 2 | 36 |
| PEM (Paeds) | 33 | 0 | 0 | 0 | 0 | 0 | 1 | 34 |
| PEM (EM) | 27 | 2 | 1 | 1 | 0 | 0 | 0 | 31 |
| Paediatrics | 6 | 0 | 0 | 0 | 0 | 0 | 0 | 6 |
| Other | 4 | 0 | 1 | 0 | 1 | 0 | 0 | 6 |
| PIC | 3 | 2 | 0 | 0 | 0 | 0 | 0 | 5 |
| **Total** | **125** | **16** | **5** | **4** | **22** | **16** | **4** | **157** |
| **Round 2** |  |  |  |  |  |  |  |  |
| Pre-hospital | 22 | 6 | 0 | 0 | 13 | 10 | 1 | 52 |
| EM | 20 | 4 | 1 | 0 | 0 | 0 | 2 | 27 |
| PEM (EM) | 21 | 2 | 1 | 0 | 0 | 0 | 0 | 24 |
| PEM (Paeds) | 23 | 0 | 0 | 0 | 0 | 0 | 1 | 24 |
| Paediatrics | 4 | 0 | 0 | 0 | 0 | 0 | 0 | 4 |
| PIC | 2 | 2 | 0 | 0 | 0 | 0 | 0 | 4 |
| Other | 4 | 0 | 0 | 0 | 0 | 0 | 0 | 4 |
| **Total** | **96** | **14** | **2** | **0** | **13** | **10** | **4** | **111** |

\*Participants were able to select multiple specialities if applicable
PEM (EM): Paediatric Emergency Medicine, Emergency Medicine background; PEM (Paeds): Paediatric Emergency Medicine, Paediatric background; EM: Emergency Medicine; PIC: Paediatric Intensive Care; ST: Speciality trainee; AS: Associate Specialist; ACP: Advanced Clinical Practitioner; ES-Paramedic: Extended Scope Paramedic

Of the 16 statements in Round 1 (R1), nine reached positive consensus; none reached negative consensus. Of the eight statements in Round 2 (R2), four reached consensus (2 positive, 2 negative); four statements did not meet the threshold for consensus. Figure 1 shows the flow of statements throughout the study.

The breakdown of responses for statements reaching consensus are presented in Table 2; those not reaching consensus after both rounds are presented in Table 3. The median score was 4 (equivalent to agree) for all statements reaching positive consensus and 2 (equivalent to disagree) for statements reaching negative consensus; the majority had an IQR of 1.

**Table 2:**
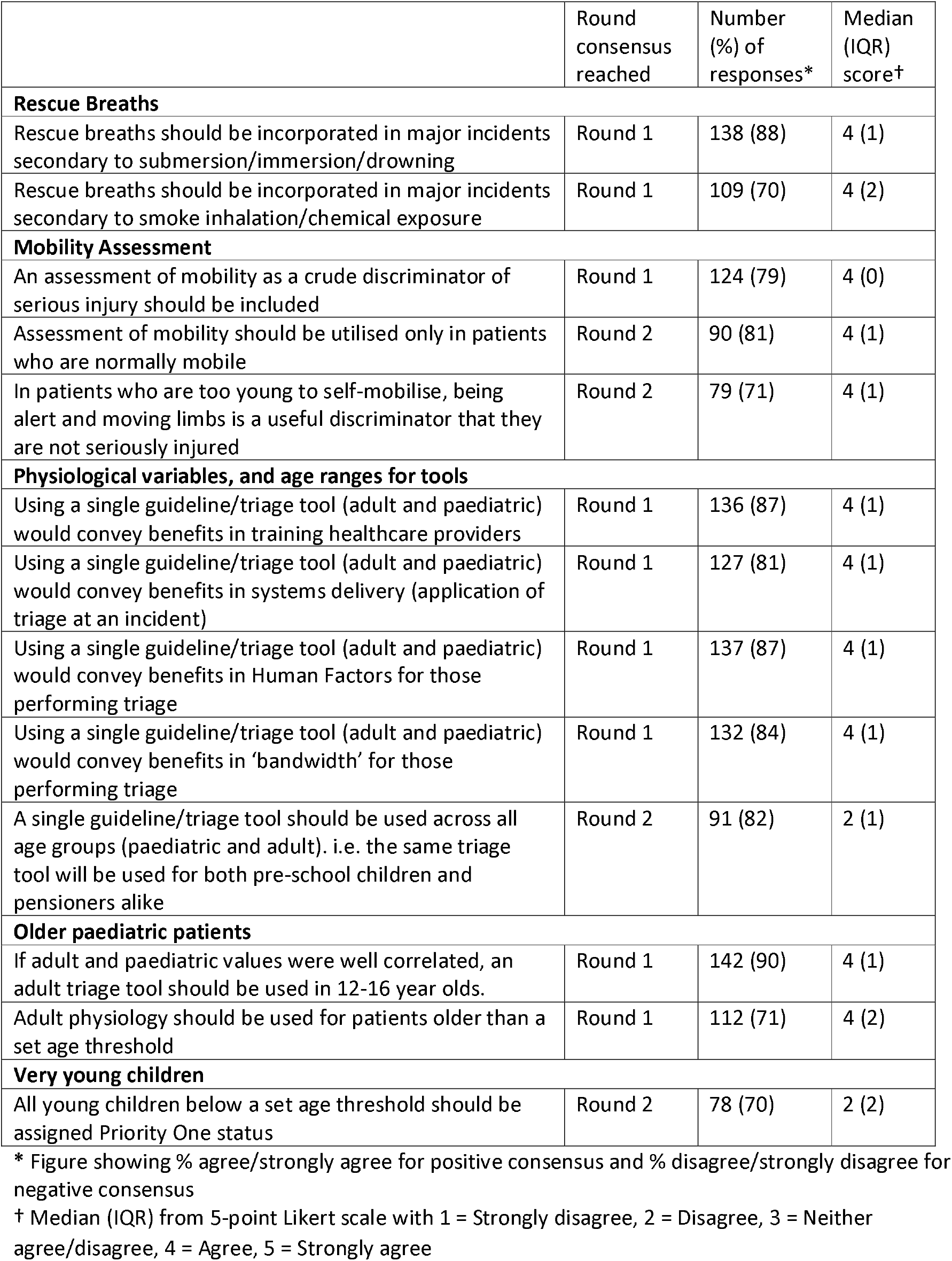
Percentages of consensus and median (IQR) scores for statements on which consensus was reached (based on ≥ 70% cut off)

|  | Round<br>consensus<br>reached | Number<br>(%) of<br>responses* | Median<br>(IQR)<br>score† |
| --- | --- | --- | --- |
| **Rescue Breaths** |  |  |  |
| Rescue breaths should be incorporated in major incidents secondary to submersion/immersion/drowning | Round 1 | 138 (88) | 4 (1) |
| Rescue breaths should be incorporated in major incidents secondary to smoke inhalation/chemical exposure | Round 1 | 109 (70) | 4 (2) |
| **Mobility Assessment** |  |  |  |
| An assessment of mobility as a crude discriminator of serious injury should be included | Round 1 | 124 (79) | 4 (0) |
| Assessment of mobility should be utilised only in patients who are normally mobile | Round 2 | 90 (81) | 4 (1) |
| In patients who are too young to self-mobilise, being alert and moving limbs is a useful discriminator that they are not seriously injured | Round 2 | 79 (71) | 4 (1) |
| **Physiological variables, and age ranges for tools** |  |  |  |
| Using a single guideline/triage tool (adult and paediatric) would convey benefits in training healthcare providers | Round 1 | 136 (87) | 4 (1) |
| Using a single guideline/triage tool (adult and paediatric) would convey benefits in systems delivery (application of triage at an incident) | Round 1 | 127 (81) | 4 (1) |
| Using a single guideline/triage tool (adult and paediatric) would convey benefits in Human Factors for those performing triage | Round 1 | 137 (87) | 4 (1) |
| Using a single guideline/triage tool (adult and paediatric) would convey benefits in 'bandwidth' for those performing triage | Round 1 | 132 (84) | 4 (1) |
| A single guideline/triage tool should be used across all age groups (paediatric and adult). i.e. the same triage tool will be used for both pre-school children and pensioners alike | Round 2 | 91 (82) | 2 (1) |
| **Older paediatric patients** |  |  |  |
| If adult and paediatric values were well correlated, an adult triage tool should be used in 12-16 year olds. | Round 1 | 142 (90) | 4 (1) |
| Adult physiology should be used for patients older than a set age threshold | Round 1 | 112 (71) | 4 (2) |
| **Very young children** |  |  |  |
| All young children below a set age threshold should be assigned Priority One status | Round 2 | 78 (70) | 2 (2) |

\* Figure showing % agree/strongly agree for positive consensus and % disagree/strongly disagree for negative consensus
† Median (IQR) from 5-point Likert scale with 1 = Strongly disagree, 2 = Disagree, 3 = Neither agree/disagree, 4 = Agree, 5 = Strongly agree

**Table 3.**
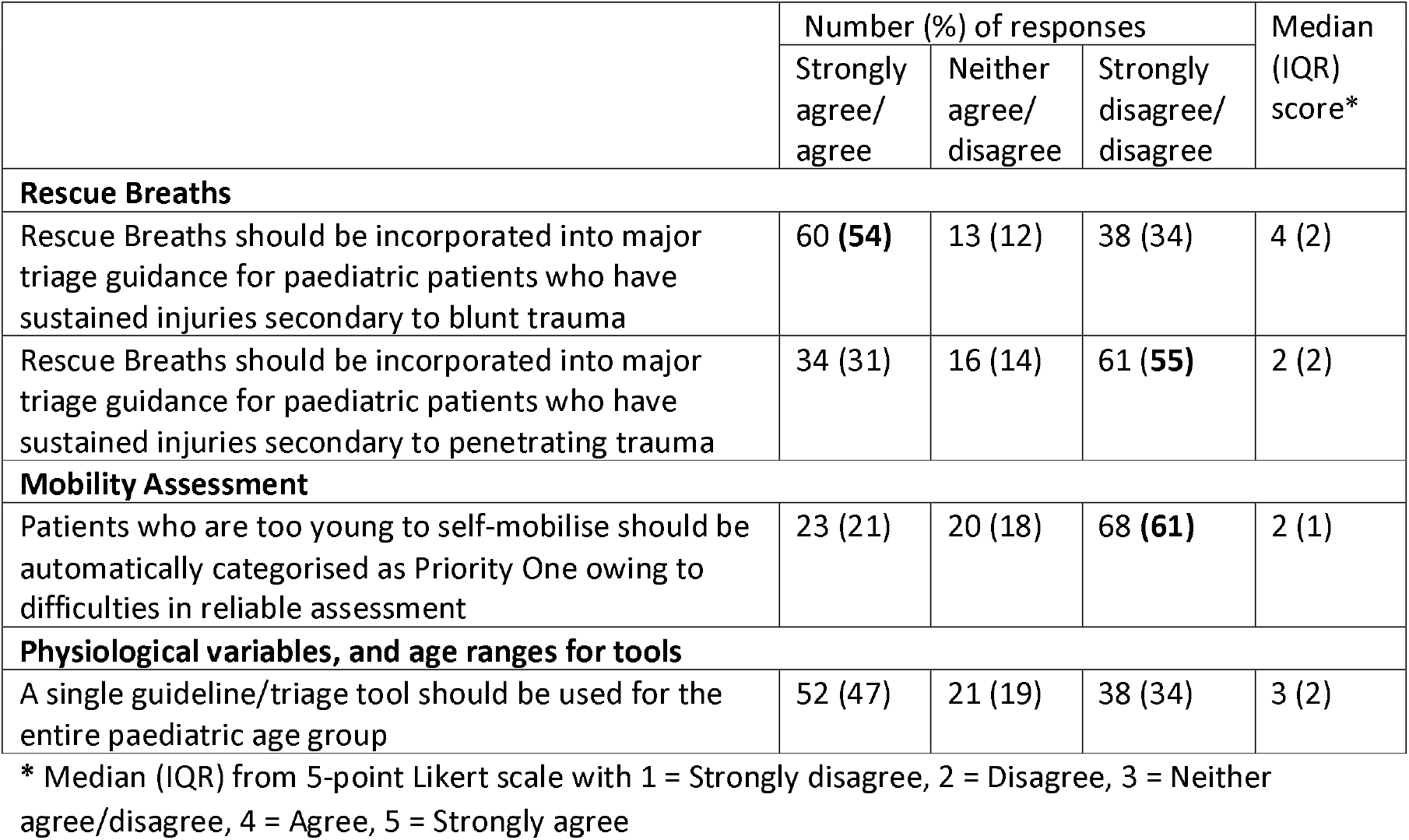
Percentages of consensus and median (IQR) score for statements that did not reach consensus (Round 2 scores)

|  | Number (%) of responses |  |  | Median (IQR) score* |
| --- | --- | --- | --- | --- |
|  | Strongly agree/ agree | Neither agree/ disagree | Strongly disagree/ disagree |  |
| Rescue Breaths |  |  |  |  |
| Rescue Breaths should be incorporated into major triage guidance for paediatric patients who have sustained injuries secondary to blunt trauma | 60 (54) | 13 (12) | 38 (34) | 4 (2) |
| Rescue Breaths should be incorporated into major triage guidance for paediatric patients who have sustained injuries secondary to penetrating trauma | 34 (31) | 16 (14) | 61 (55) | 2 (2) |
| Mobility Assessment |  |  |  |  |
| Patients who are too young to self-mobilise should be automatically categorised as Priority One owing to difficulties in reliable assessment | 23 (21) | 20 (18) | 68 (61) | 2 (1) |
| Physiological variables, and age ranges for tools |  |  |  |  |
| A single guideline/triage tool should be used for the entire paediatric age group | 52 (47) | 21 (19) | 38 (34) | 3 (2) |

\* Median (IQR) from 5-point Likert scale with 1 = Strongly disagree, 2 = Disagree, 3 = Neither agree/disagree, 4 = Agree, 5 = Strongly agree

### Statements reaching consensus agreement

#### Rescue Breaths

Consensus was reached in Round 1 that rescue breaths should be used in major incidents secondary to submersion/immersion/drowning (88%), and smoke inhalation/chemical exposure (70%).

#### Mobility Assessment

It was agreed that an assessment of mobility should be included as a crude discriminator for serious injury (R1, 79%) and that this assessment should only be used in patients who are normally mobile (R2, 81%). In patients too young to self-mobilise, being alert and moving limbs was felt to be useful in determining absence of serious injury (R2, 71%).

#### Physiological variables, and age ranges for tools

Use of a single MIT tool was felt to convey benefits in training healthcare providers (R1, 87%), delivery (application of a tool in a major incident; R1, 81%), human factors (R1, 87%), and “bandwidth” (R1, 84%) for those performing triage. However, in contrast with these findings, the consensus of participants was that a single tool should not be used across all age groups. (R2, 82%)

#### Older paediatric patients

It was agreed that if adult and paediatric physiological values are well correlated, an adult triage tool should be used in 12-16 year-olds (R1, 90%). Adult physiology was also agreed to be valid for use in patients above an age threshold (R1, 71%) with the majority (92%) suggesting >12 years.

#### Very young children

Consensus was reached that children younger than a given age threshold should not automatically be assigned P1 status. (R2, 70%)

### Statements not reaching consensus agreement

For statements on which consensus was not reached (Table 3), predominantly positive, negative, or neutral responses are shown in bold. Responses were reviewed in line with the clinical background of respondents to determine whether there were any marked differences between professional groups. In regards to rescue breaths in blunt trauma, consultants were more likely to agree that they had a role in MIT (62%), whilst paramedics were more likely to disagree (52%). Incongruity was less marked between speciality groups and grades for other statements not reaching consensus (Supplementary Table 1).

## DISCUSSION

In this modified Delphi study, we have achieved consensus on thirteen statements regarding paediatric MIT, from a large number and range of healthcare professionals likely to encounter a paediatric major incident. Agreement was reached that rescue breaths should be included when the mechanism related to hypoxic or asphyxiating injury, and that adult physiology thresholds should be used for older children. An assessment of mobility as a discriminator for serious injury should be used in children who are normally mobile, and for younger non-mobile patients, being alert and moving limbs was felt to be an appropriate surrogate. Despite benefits of a single MIT tool being appreciated by participants, consensus was reached that a single tool should not be used across all ages.

Whilst positive consensus was reached supporting use of a single MIT tool from the perspective of training and systems delivery, we derived negative consensus regarding clinical implementation of a single tool across all ages. This was despite participants being provided with details of a recently published study as contextual information, which demonstrated that the MPTT-24 outperformed existing paediatric MIT tools at identifying patients in need of life-saving interventions using trauma registry data.^10^ With no qualitative feedback from Delphi participants on this area, we can only hypothesise that clinicians felt uncomfortable adopting a completely new approach to paediatric MIT (i.e. not using a bespoke paediatric tool). Specific to the paediatric age range, consensus was not reached as to whether a single MIT tool should be used across all ages. This uncertainty may represent mixed opinion on the trade-off between incorporating physiological variables against ease of use. For example, the Sheffield Paediatric Triage Tool has physiological thresholds which vary with age, which increases sensitivity within age bands; however, these require greater cognitive input by operators, rendering the tool more complicated and potentially reducing its effectiveness in the pre-hospital setting of a major incident.

Consensus could not be reached on incorporating rescue breaths for blunt or penetrating trauma. For blunt trauma, participant responses showed a trend against their use, but this did not meet the pre-specified criteria for consensus. Between respondent groups, there was disagreement between consultants and paramedics, which was not replicated in the context of penetrating trauma. Whilst rescue breaths were felt to be appropriate in mechanisms associated with hypoxia or asphyxiation (e.g. smoke inhalation or immersion), and are clinically appropriate in single patient incidents, they may be impractical in the major incident setting for a variety of reasons. Firstly, if multiple patients require rescue breaths this will require multiple bag-valve-masks (and potentially airway adjuncts) of varying sizes. Secondly, if being performed as a sole practitioner (which may well be the case initially), achieving successful ventilation can be difficult^12^. Lastly, and of particular significance in large marauding terrorist incidents, there will be an increased time allocation for patients receiving rescue breaths, with a resultant deleterious effect on patients who are yet to be triaged (who may for example have catastrophic haemorrhage requiring immediate intervention).

Recent evidence has demonstrated that younger children (aged under 2 years) have the highest mortality and need for life-saving interventions in a trauma registry population.^10^ As this cohort is potentially difficult to assess on scene, one suggested strategy has been that all such children be automatically categorised as P1 to reduce cognitive burden for operators.^11^ However despite this evidence being presented, negative consensus was reached in regards to this approach. Similarly, although consensus was not reached, two-thirds of participants stated that non-mobile children should not be automatically categorised as P1. This may be due to concerns surrounding unnecessary prioritised conveyance, potentially to the detriment of more seriously injured patients pre- and in-hospital. Given this offset between opinion and evidence of risk, this area is worthy of further study. In contrast, for normally non-mobile children, consensus was reached that being alert and moving limbs is a useful discriminator for exclusion of serious injury, in keeping with current practice.

## Limitations

We acknowledge the limitations associated with our study. Firstly, only healthcare professionals working in the UK and Ireland were included, and our findings may not be generalisable to other settings (for example those in low resource countries, or with different frequency and characteristics of paediatric major incidents). Secondly, whilst the methodology allowed for maximal uptake engagement of relevant professionals, we are unable to determine a ‘denominator’ for our subject matter expert response, and we acknowledge the possibility of selection bias. However, we believe this is offset by a high completion rate across both rounds (71%). Thirdly, exclusion of incomplete responses may have influenced whether consensus was reached for statements; however, in a secondary analysis which included partial responses, there was no change in our findings. Finally, contextual evidence provided to participants was from analysis of trauma registry data, rather than major incident datasets. No such datasets exist, and the unpredictable nature of major incidents makes prospective research difficult and potentially unethical. The use of trauma registry data to develop and validate MIT tools has therefore previously been used successfully in development of adult MIT tools.

## Conclusion

This Delphi study has established consensus among a large group of subject matter experts on several key elements of paediatric MIT, which will be of use to those involved in planning and preparing the response to a major incident involving children. We would encourage further work to develop a triage tool that can be implemented based on emerging and ongoing research in addition to the consensus opinion demonstrated in this study.

## Data Availability

All data produced in the present study are available upon reasonable request to the authors

## Statements

### Ethical Approval

No formal research ethics approval was required for this study as it was a survey of health professionals identified via networks. Participation was deemed as consent.

### Clinical Trial Registration

Not applicable.

## Acknowledgements

We would like to thank the following participants who completed both rounds of the Delphi and who consented for us to provide their names:

R James, O Cubitt, M Tehan, S Amps, C Gray, C McGahan, R Bayliss, W Thomson, M Tunnicliff, K Challen, A Price, K Allen, A Baron, N Malcolm, M Booker, JA Maney, JA Cowper, A Cowper, C Brown, L Draper, M England, K Charlick, L Bagshaw, H Procter, N Dole, M Stewart, S Graham, J Tooley, M Lyttle, A Frampton, S Milsom, R Moy, M Tolhurst-Cleaver, M Harrison, C Fitchett, D Bramley, E Walton, C Stutchfield, J Loughrey, S Keers, C Gough, A Bradley, S Morton, S Hingley, J McCormack, C O’Connell, L Melody, J Bayreuther, A Maddock, T Hurst, V Brooker, H Stewart, S McKenna, R Moss, B Taylor, H Mekki, S Wilson, S Mullen, C Nunn, J Hudge, C Sutton, E Day, J Bailey, C Dieppe, A Holdstock, C Johnson, A Alcock, R Cooper, S Stibbards, T James, J Surridge, J Raitt, P Reeve, N Creasey, M Barrett, H Murch, E Saunders, J Browning, T Coats, N Howarth, J Stirling, T Bolger, I Lewins, A Crawford, M Little, D Broomfield, A Kidd, L Reid, N Adams, G Hann, H Rutkowska, P Leonard, J Watson, C Hill, K Botsford, M McGlone, L McCreddan, J Plumb, R Harding

## Author contributions

JV and ML conceived the study idea, with survey content designed by JV, JES, PC, ML. The study was conducted by JV and ML. JV, SB, ML conducted the analysis and interpretation. SB wrote the initial draft of the manuscript. All authors contributed with critical revisions to the manuscript and approved the final manuscript. JV takes responsibility for the manuscript as a whole.

## Patient and public involvement

Patients and/or the public were not involved in the design, or conduct, or reporting or dissemination plans of this research.

